# Tocilizumab is associated with reduced risk of ICU admission and mortality in patients with SARS-CoV-2 infection

**DOI:** 10.1101/2020.06.05.20113738

**Authors:** Estela Moreno-García, Verónica Rico, Laia Albiach, Daiana Agüero, Juan Ambrosioni, Marta Bodro, Celia Cardozo, Mariana Chumbita, Lorena De la Mora, Nicole García-Pouton, Carolina Garcia-Vidal, Ana González-Cordón, Marta Hernández-Meneses, Alexy Inciarte, Montse Laguno, Lorna Leal, Laura Linares, Irene Macaya, Fernanda Meira, Josep Mensa, Antonio Moreno, Laura Morata, Pedro Puerta-Alcalde, Jhon Rojas, Montse Solá, Berta Torres, Manuel Torres, Adrià Tomé, Pedro Castro, Sara Fernández, Josep Maria Nicolás, Alex Almuedo-Riera, Jose Muñoz, Mariana Jose Fernandez-Pittol, Maria Angeles Marcos, Dolors Soy, José Antonio Martínez, Felipe García, Alex Soriano

**Author notes:** both authors contributed equally to this manuscript. **Corresponding author:** Alex Soriano, M.D. Ph.D., Department of Infectious Diseases, Hospital Clínic of Barcelona., Carrer de Villarroel 170, 08036, Barcelona, Spain.

## Abstract

**Background:** In some patients the immune response triggered by SARS-CoV-2 is unbalanced, presenting an acute respiratory distress syndrome which in many cases requires intensive care unit (ICU) admission. The limitation of ICU beds has been one of the major burdens in the management worldwide; therefore, clinical strategies to avoid ICU admission are needed.

**Objective:** We aimed to describe the influence of tocilizumab on the need of transfer to ICU or death in non-critically ill patients.

**Methods:** A retrospective study of 171 patients with SARS-CoV-2 infection that did not qualify as requiring transfer to ICU during the first 24h after admission to a conventional ward, were included. The criteria to receive tocilizumab was radiological impairment, oxygen demand or an increasing of inflammatory parameters, however, the ultimate decision was left to the attending physician judgement. The primary outcome was the need of ICU admission or death whichever came first.

**Results:** 77 patients received tocilizumab and 94 did not. The tocilizumab group had less ICU admissions (10.3% vs. 27.6%, *P*= 0.005) and need of invasive ventilation (0 vs 13.8%, *P*=0.001). In multivariable analysis, tocilizumab remained as a protective variable (OR: 0.03, CI 95%: 0.007-0·1, *P*=0.0001) of ICU admission or death.

**Conclusion:** Tocilizumab in the early stages of the inflammatory flare, could reduce ICU admissions and mechanical ventilation use. The mortality rate of 10.3% among patients receiving tocilizumab appears to be lower than other reports.

**Clinical implication:** Our results suggest that tocilizumab administered to non-critically ill patients could reduce ICU admissions and mortality.

**Capsule summary:** Tocilizumab administered to non-critically ill patients with SARS-CoV-2 infection in the early stages of the inflammatory flare, could reduce an important number of ICU admissions and mechanical ventilation use.

## Introduction

Infection by Coronavirus 2 (SARS-CoV-2) emerged in December 2019 in Wuhan and rapidly spread around the world. SARS-CoV-2 pneumonia evolves in 2 different phases, the first one is characterized by a high viral replication and classical symptoms of a respiratory virus infection, including fever, malaise, myalgia, and cough (1). About 80% of the patients control the infection within a week but 20% of them, after the first 7-8 days, develop a severe respiratory failure fulfilling the definition of acute respiratory distress syndrome (ARDS) with many requiring intensive care management (2). During the second week, the blood tests reveal lymphopenia, and high levels of C-reactive protein (CRP), ferritin, and D-dimer values (1), all related with the activity of different cytokines (IL-1beta, IL-2, IL-6, IL-8, IL-17, IFN-gamma or TNF-alpha) (3). Therefore, the main therapeutic objective during the first week of treatment is to stop the viral replication while in the second week blocking the tissue damage induced by the cytokine storm is paramount (4).

In agreement with the immunopathogenesis, it has been proposed to treat patients during the inflammatory flare with IL-6 inhibitors (5,6). The first description included 21 patients admitted to a Chinese hospital who received tocilizumab, a recombinant humanized anti-IL-6 receptor monoclonal antibody. In a few days, symptoms improved remarkably, 75% had lowered their oxygen intake and no patient died (7). More recently, Sciascia and colleagues, have reported the results of a single-arm multicentre study on off-label use of tocilizumab conducted in 63 Italian patients with a severe SARS-CoV-2 infection. The administration of tocilizumab within 6 first days of admission was associated with an increased likelihood of survival (HR 2.2 95% CI 1.3-6.7, p<0.05) (8).

On the other hand, 21 Italian patients received siltuximab, a chimeric monoclonal antibody that binds to and blocks the effect of IL-6. The outcome showed that 33% (7/21) of patients experienced an improvement in their condition with a reduced need for ventilation, 43% (9/21) remained stable, and 24% (5/21) worsen and required intubation. However, this article is included in a repository so not yet peer reviewed (9). The main objective of the present article is to describe the putative influence of tocilizumab on the prognosis, defined as eventual need of transfer to the ICU or death, of 171 non-critically ill patients.

## Methods

From February 19^th^ to April 16^th^ patients with respiratory symptoms and radiological evidence of pneumonia (uni or bilateral interstitial infiltrates) and those with respiratory symptoms without pneumonia but with co-morbidity (hypertension, diabetes mellitus, cancer, chronic liver diseases, chronic obstructive pulmonary disease or immunosuppression) were admitted to Hospital Clínic of Barcelona in the context of SARS-CoV-2 pandemic. Definitive diagnostic was stablished by a positive polymerase chain reaction (PCR) from a nasopharyngeal swab during the first two weeks of pandemic but once the prevalence of positive tests was >70%, the diagnosis was based on clinical criteria. Clinical criteria for defining a case of SARS-CoV-2 were the presence of respiratory symptoms with uni or bilateral interstitial infiltrate in the chest-X ray without evidence of other potential causes (e.g. heart failure). During the study period, 171 patients that did not qualify as requiring transfer to the ICU during the first 24h after admission to a conventional hospital ward, were included.

For ARDS, the Berlin definition (10) was applied. When PaO_2_ was not available, SpO_2_/FiO_2_ ≤ 315 suggested ARDS (including in non-ventilated patients) (11).

The standard protocol included antiviral treatment that consisted of lopinavir/ritonavir 400/100 mg BID for 7-14 days plus hydroxychloroquine 400 mg/12h on the first day, followed by 200 mg/12h for the next 4 days. From the 18^th^ of March onwards, azithromycin 500 mg the first day and 250 mg/24h for 4 additional days was added to the regimen. In addition, a clinical trial with remdesivir was enrolling patients in our institution during the study period. All patients with risk factors for thrombosis received prophylactic doses of low molecular weight heparin (12). Intravenous methylprednisolone was recommended for patients with disease progression to ARDS. The local protocol suggested the use of tocilizumab for patients with pneumonia, progressive respiratory failure (increasing fraction of inspired oxygen) and CRP ≥ 8 mg/dL or ferritin ≥800 ng/mL or lymphocyte count < 800 cells/mm^3^. The dose was 400 mg/24h iv for patients with ≤75 kg and 600 mg/24h iv for those with >75 kg with the possibility to repeat the dose every 12h up to 3 doses in case of only partial response. However, due to the lack of evidence to support its efficacy, the ultimate decision about using tocilizumab was left to the judgement of the attending physician.

Patients with severe comorbidity and a life expectancy <6 months were considered no tributary of advanced life support (ALS). The outcome variable was a composite of the need of ICU admission or death whichever came first. The last revision of medical charts was April 26^th^.

The Institutional Ethics Committee of the Hospital Clínic of Barcelona approved the study and due to the nature of retrospective chart review, waived the need for inform consent from individual patients (Comité Ètic d’Investigació Clínica; HCB/2020/0273).

### Statistical analysis

Categorical variables were described using the absolute number and percentage and continuous variables using the mean and standard deviation (SD). Categorical variables were compared using a Chi-squared test or Fisher exact test when necessary, and means by using the Student-t test. For multivariable analysis, variables with a *P*-value ≤ 0.2 in the univariable analysis were subjected to further selection by using a backward logistic regression procedure. Interactions between variables were explored. In order to reduce the effect of selection bias, we estimate the propensity score (PS) to receive tocilizumab as the predicted probability from a logistic regression model using tocilizumab as the dependent variable. The PS was included in the multivariable analysis of the main outcome. The calibration of the model was assessed by means of the Hosmer-Lemeshow goodness-of-fit test. Statistical significance was defined as a two-tailed *P* value <0.05. The analysis was performed in SPSS version 23 (SPSS Inc., Chicago, IL).

## Results

The cohort included 171 patients, of whom 77 received tocilizumab while staying in a conventional ward and 94 did not, with a mean (SD) age of 61.5 (12.4) and 61.4 (16) years, respectively. The proportion of males and the main comorbidities were similar between both groups (Table 1). Patients in the tocilizumab group had more frequently fever, pneumonia (interstitial infiltrate) and at day 1 they needed more often oxygen therapy. C-reactive protein levels were significantly higher in the tocilizumab group (9.7 mg/dL vs. 7.5 mg/dL, *P*=0.04) but other biological parameters were similar in both groups. During patients’ stay in a conventional ward, corticosteroid therapy was more frequently administered in the tocilizumab group (50.6% vs. 27.7%, *P*=0.002). A total of 26 patients were not candidates to ALS, 10 (12.9%) in the tocilizumab group and 16 (17%) among controls. The mean (SD) time from symptoms onset to hospital admission in tocilizumab group was 6.5 (3.3) days while it was 5 (6.5) days in the control group. The outcome of both groups, with >90% of patients discharged alive or dead, showed that patients in the tocilizumab group had significantly less ICU admissions (10.3% vs. 27.6%, *P*= 0.005) and less need of invasive ventilation (0 vs 13.8%, *P*=0.001) (Figure 1). The univariable analysis of our composite outcome (ICU admission or death whichever came first) showed that comorbidities (hypertension, heart diseases and lymphoma), the need of oxygen at day 1, a CRP > 16 mg/dL and the development of cardiovascular, renal or respiratory (ARDS, invasive ventilation) complications were significantly associated with the primary outcome. In contrast, tocilizumab was the only one protective variable (Table 2). In the multivariable analysis, including the PS estimate to receive tocilizumab as a potential confounder, tocilizumab remained as a strong protective variable (OR: 0.03, CI 95%: 0.007-0.1, *P*=0.0001) of ICU admission or death (Table 3).

**Table 1.** Characteristics and outcome of patients that received or did not received tocilizumab in a conventional ward.

| **Variables** | **Tocilizumab group**<br>**(N=77)** | **Control group**<br>**(N=94)** | **P - value** |
| --- | --- | --- | --- |
| Mean (SD) age in years | 61.5 (12.4) | 61.4 (16.0) | 0.957 |
| Age > 62 years old (%) | 40 (52) | 52 (55.3) | 0.660 |
| Male (%) | 53 (68.8) | 59 (62.7) | 0.406 |
| **Comorbidities (%)** |  |  |  |
| Hypertension | 35 (45.4) | 43 (45.7) | 0.960 |
| Heart diseases | 12 (15.5) | 21 (22.3) | 0.265 |
| Chronic respiratory disease | 8 (10.3) | 12 (12.7) | 0.630 |
| Diabetes Mellitus | 12 (15.6) | 14 (15) | 0.900 |
| Mean (SD) days from symptoms onset to admission | 6.5 (3.3) | 5 (6.5) | 0.061 |
| **Initial characteristics (%)** |  |  |  |
| Fever | 86 (98.7) | 80 (85) | 0.002 |
| Dyspnea | 33 (43) | 47 (50) | 0.352 |
| Cough | 64 (83) | 70 (74.5) | 0.172 |
| Normal chest x-ray at admission | 3 (4) | 14 (15) | 0.017 |
| Need of oxygen therapy at day 1 | 56 (72.7) | 50 (53.8) | 0.011 |
| Positive PCR from a nasal swab | 68 (88.3) | 82 (87.2) | 0.831 |
| **Laboratory at admission mean (SD)** |  |  |  |
| D-dimer (ng/mL)<sup>1</sup> | 918.6 (1354.8) | 1503.9 (2175.4) | 0.100 |
| Lymphocytes count (cell/mm<sup>3</sup>) | 878.9 (452.8) | 910.1 (534.6) | 0.686 |
| C-Reactive protein (mg/dL)<sup>2</sup> | 9.7 (7.4) | 7.5 (5.7) | 0.044 |
| Serum ferritin (ng/dL)<sup>3</sup> | 867.8 (871) | 904.1 (809.9) | 0.842 |
| ARDS at any given time (%) | 24 (31.1) | 26 (27.6) | 0.616 |
| **Treatments received (%)** |  |  |  |
| Antiviral agents<sup>4</sup> | 77 (100) | 91(96.8) | 0.164 |
| Steroid prior ICU admission | 39 (50.6) | 26 (27.7) | 0.002 |
| Not candidate to ALS (%) | 10 (12.9) | 16 (17) | 0.465 |
| Mean (SD) days of follow up | 11.2 (6.2) | 14.7 (10.6) | 0.027 |
| **Outcomes (%)** |  |  |  |
| Need of ICU | 8 (10.3) | 26 (27.6) | 0.005 |
| Need of no invasive MV | 3 (3.9) | 1 (1) | 0.198 |
| Need of invasive MV | - | 13 (13.8) | 0.001 |
| Extubation | - | 9 (9.6) | - |
| Discharge from ICU | 5 (6.5) | 21 (22.3) | - |
| Hospital discharged | 65 (84.4) | 71 (75.5) | 0.152 |
| Still in the hospital | 4 (5.2) | 6 (6.4) | 0.503 |
| **Mortality (%)** |  |  |  |
| Global mortality | 8 (10.3) | 17 (18) | 0.156 |
| Mortality in: |  |  |  |
| Not candidates to ALS | 6 (60) | 12 (75) | 0.420 |
| Candidates to ALS | 2 (3) | 5 (6.4) | 0.337 |

PCR, polymerase chain reaction. ADRS, adult distress respiratory syndrome. ICU, intensive care unit.
ALS, advanced life support. MV, mechanical ventilation.
<sup>1</sup> Measured in 110 patients.
<sup>2</sup> Measured in 168 patients.
<sup>3</sup> Measured in 86 patients.
<sup>4</sup> see material and methods for antivirals used in our protocol.

**Figure 1.**
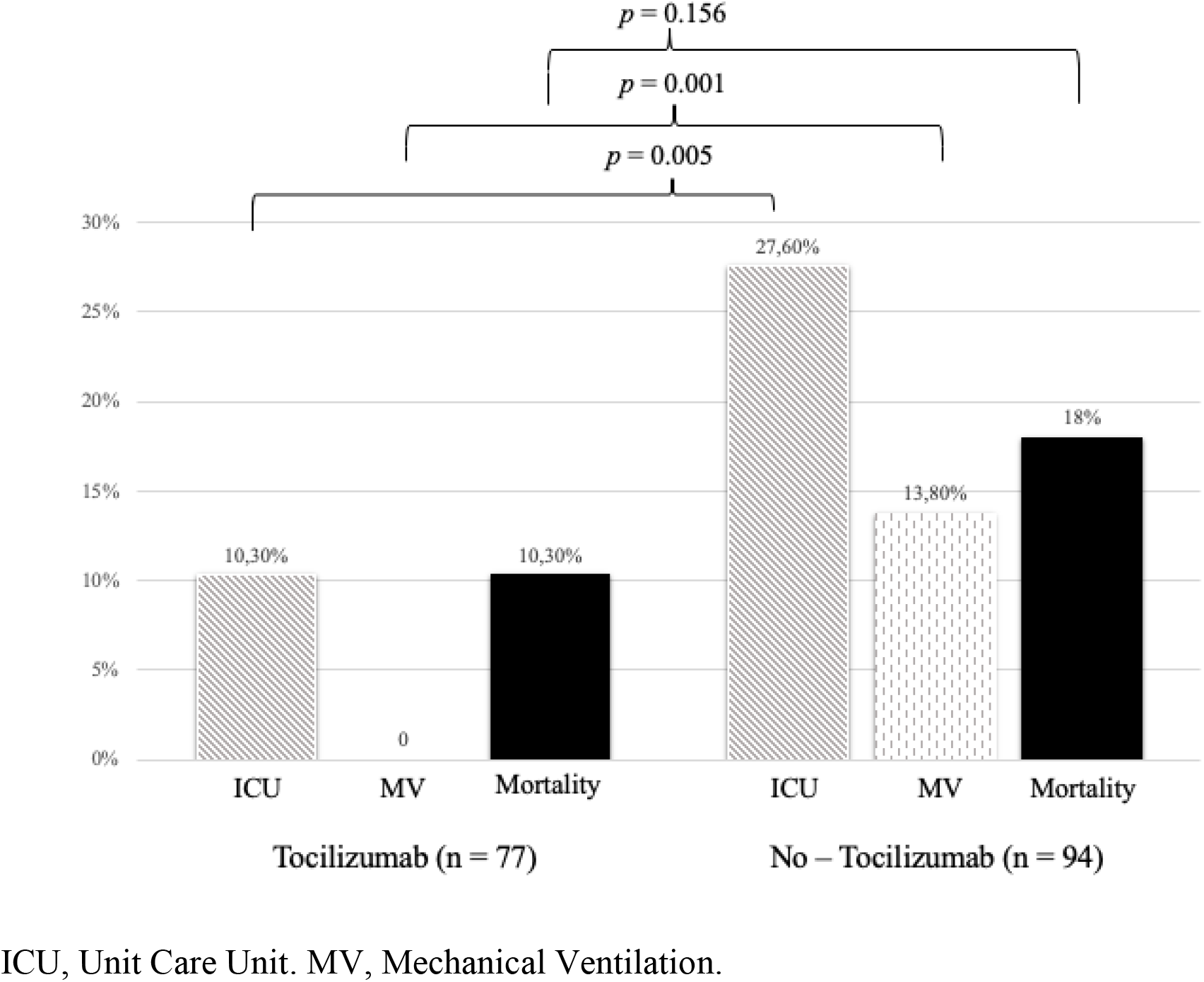
Differences between tocilizumab group and no-tocilizumab group in terms of ICU admission, need of mechanical ventilation and mortality.

**Table 2.** Variables associated with ICU admission and/or death whichever came first.

| Variables | No ICU admission<br>and/or death, N=121 | ICU admission or<br>death, N=50 | P - value |
| --- | --- | --- | --- |
| Age >62 years (%) | 57 (47) | 35 (70) | 0.006 |
| Male sex (%) | 77 (63.6) | 35(70) | 0.426 |
| Mean (SD) follow-up, days | 12 (8.347) | 16.6 (9.858) | 0.006 |
| **Comorbidities (%)** | 98 (81) | 48 (96) | 0.012 |
| Hypertension | 49 (40.5) | 29 (58) | 0.037 |
| Diabetes Mellitus | 20 (16.5) | 6 (12) | 0.453 |
| Heart diseases | 17 (14) | 16 (32) | 0.007 |
| Chronic respiratory disease | 11 (9) | 9 (18) | 0.099 |
| Neoplasia | 11(9) | 6(12) | 0.580 |
| Dyslipemia | 8 (6.6) | 6 (12) | 0.356 |
| Lymphoma | 2 (1.7) | 5 (10) | 0.012 |
| Solid organ transplantation | 5 (4) | 3 (6) | 0.693 |
| Human Immunodeficiency Virus | 1(0.8) | - | 1 |
| Mean (SD) days from symptoms onset to admission | 5.98 (6.124) | 4.86 (3.084) | 0.223 |
| **Initial characteristics (%)** |  |  |  |
| Fever | 112(92.6) | 44 (88) | 0.337 |
| Dyspnoea | 59 (48.8) | 21 (42) | 0.420 |
| Cough | 98 (81) | 36 (72) | 0.222 |
| Normal chest x-ray at admission | 12 (10) | 5 (10) | 0.987 |
| Need of oxygen therapy at day 1 | 68 (56.7) | 38 (76) | 0.018 |
| Positive PCR from nasal swab | 105 (86.8) | 45 (90) | 0.559 |
| **Laboratory at admission (%)** |  |  |  |
| Lymphocytes count<sup>1</sup> <700 cell/mm<sup>3</sup> | 36 (29.8) | 18 (36) | 0.424 |
| C-Reactive protein<sup>2</sup> >9 mg/dL | 39 (32.5) | 20 (41.7) | 0.261 |
| C-Reactive protein<sup>2</sup> >16 mg/dL | 23 (19) | 25 (50) | 0.0001 |
| **Treatments received (%)** |  |  |  |
| Antiviral agents | 118 (97.5) | 50 (100) | 0.261 |
| Steroids prior ICU admission | 44 (36.4) | 21 (42) | 0.490 |
| Tocilizumab | 65 (53.7) | 12 (24) | <0.0001 |
| **Complications (%)** |  |  |  |
| Cardiovascular | 5 (4) | 13 (7.6) | 0.012 |
| ARDS in the conventional ward | 14 (11.6) | 30 (60) | 0.0001 |
| Acute Kidney Injury | 3 (1.8) | 12 (24) | 0.0001 |
| Invasive ventilation | - | 13 (26) | 0.0001 |
| **Not candidate to ALS** | 8 (6.6) | 18 (36) | 0.0001 |

PCR, polymerase chain reaction. ICU, intensive care unit. ADRS, adult distress respiratory syndrome.
ALS, advanced life support.
<sup>1</sup> Measured in 110 patients.
<sup>2</sup> Measured in 168 patients.
<sup>3</sup> See material and methods section for antivirals used in our protocol.

**Table 3.**
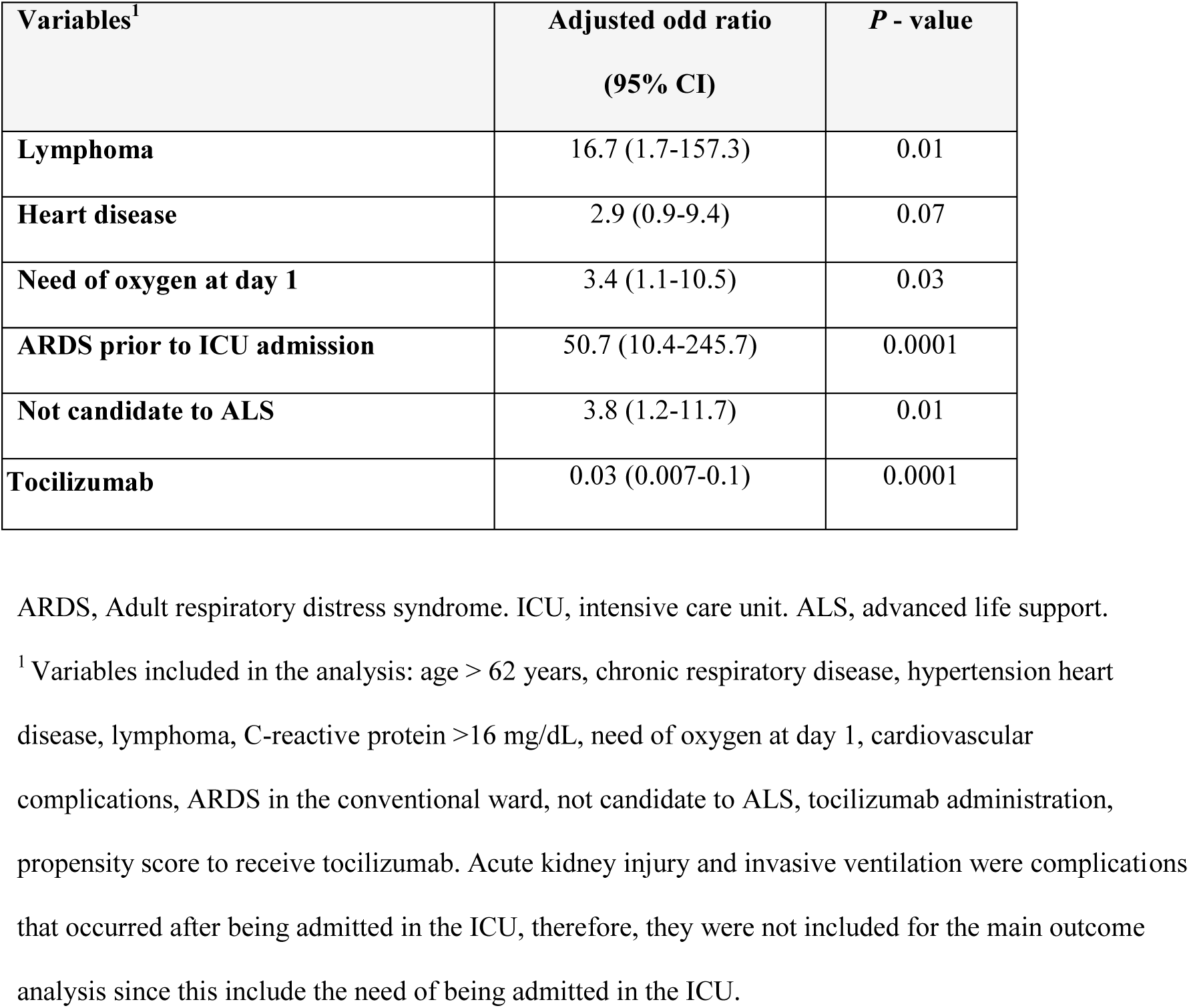
Variables significantly associated with the risk of being admitted in the ICU and/or death in the multivariable analysis.

## Discussion

Monoclonal antibodies directed against key inflammatory cytokines represent a class of potential adjunctive therapies for SARS-CoV-2 infected patients. The rationale for their use is that the underlying pathophysiology of significant lung damage is caused by a cytokine storm being IL-6 one of the main drivers. Therefore, monoclonal antibodies against IL-6 or its receptor could theoretically improve clinical outcomes mainly by reducing the need of ICU admission and consequently the associated mortality. Tocilizumab, a monoclonal antibody IL-6 receptor antagonist, was administered to 77 patients admitted to a conventional ward in our hospital and the outcome was compared with 94 patients also admitted in a conventional ward during the same period of time that did not receive tocilizumab. Although this study was not randomized, the characteristics of both groups did not differ in terms of demographics and comorbidities Moreover, the tocilizumab group had more severe infection (pneumonia, need of oxygen at day 1 or higher CRP). Furthermore, all the patients were evaluated during the same period of time so the same criteria for being transferred to the ICU was applied. After adjusting for potential confounders, including the PS for receiving tocilizumab, the multivariable analysis revealed that tocilizumab was an independent factor associated with a reduction in the need of ICU admission and death. The need of ICU in the tocilizumab group was almost 3 times lower (10.3% vs. 27.6%) than in controls and it was lower than the one reported in Wuhan hospitals (26%) (1,13) or more recently in New York (14%) (14). The availability of ICU beds is critical for the management of patients that develop a severe ARDS in few hours, therefore, reducing the need of ICU beds using tocilizumab impacted directly not only on the outcome of patients that received the treatment but also of those that not receiving tocilizumab or arriving too late in a critically ill condition had more chances of being admitted in the ICU. In line with this, the mortality of our cohort, including patients not candidates to ALS, was 14.2% which seems lower than that showed in previous reports, regularly >20% (1,13,15). Indeed, the mortality rate of patients in the tocilizumab group was 10% that is comparable to the one recently reported with remdesivir (15) in 53 patients (13%). Although from the beginning of the pandemic tocilizumab was recommended in the general protocol, the heterogenicity of its prescription could be explained by the lack of clinical randomized trials supporting its usefulness.

Our results suggest that tocilizumab should be administered in early phases of the inflammatory flare. It is reasonable to hypothesize that other strategies directed to inhibit other specific inflammatory pathways (including IL-1 with anakinra or INF-gamma with JAK inhibitors), or a broad-spectrum inhibition with steroids with or without therapeutic strategies to reduce the pro-coagulant status, could be also effective (16–18). On the other hand, although in non-severe cases after one week from symptoms onset the viral viability is significantly reduced, there is data supporting the continuous viral replication in severe cases (19) that could be the trigger for the inflammatory flare and its maintenance. In addition, different antiviral strategies (antiviral agents, plasma from convalescent patients) have demonstrated benefit in severe patients, even >1 week after symptoms onset (15, 20–21). Accordingly, we consider that antiviral agents should be associated with immunomodulators (22), and this explains why almost 100% of our cohort received it.

In conclusion, while waiting for the definitive results from clinical trials with tocilizumab (NCT04310228, ChiCTR200002976), our findings support that the administration of tocilizumab in the early stages of the inflammatory flare, particularly before the need of ICU admission, is more convenient and could potentially avoid an important number of ICU admissions and mechanical ventilation use. Consequently, the mortality rate of 10.3% among patients receiving tocilizumab appears to be lower than that described by others in previously published series (13,15,23). However, this is a non-randomized study and, therefore, the results should be interpreted with caution.

## Data Availability

The data of this study will be available upon request to the corresponding author.

## Acknowledgements

Hospital Clinic of Barcelona COVID-19 Research Group:

Infectious Diseases’ Research Group:

Blanco JL, Mallolas J, Martínez E, Martínez M, Miró JM, Moreno A, and all the staff members.

Medical Intensive Care Unit:

Adrian Téllez, and all the staff members.

Department of International Health:

Daniel Camprubi Ferrer, Maria Teresa de Alba, Marc Fernandez, Elisabet Ferrer, Berta Grau, Helena Marti, Magdalena Muelas, Maria Jesus Pinazo, Natalia Rodríguez, Montserrat Roldan, Carme Subira, Isabel Vera, Nana Williams, Alex Almuedo-Riera, Jose Muñoz, and all the staff members.

Department of Internal Medicine:

Aldea A, Camafort M, Calvo J, Capdevila A, Cardellach F, Carbonell I, Coloma E, Foncillas A, Estruch R, Feliu M, Fernández-Solá J, Fuertes I, Gabara C, Grafia I, Ladino A, López-Alfaro R, López-Soto A, Masanés F, Matas A, Navarro M, Marco-Hernández J, Miguel L, Milisenda J, Moreno P, Naval J, Nicolás D, Oberoi H, Padrosa J, Prieto-González S, Pellicé M, Ribot J, Rodríguez-Núnez O, Sacanella E, Seguí F, Sierra C, Ugarte A, Ventosa H, Zamora-Martínez C, and all the staff members.

Department of Microbiology:

Almela M, Alvarez M, Bosch J, Casals C, Costa J, Cuesta G, Fidalgo B, Gonzàlez J, Hurtado JC, Marco F, Martínez M, Mosquera M, Narvaez S, Pitart C, Rubio E, Vergara A, Valls ME, Vila J, Zboromyrska Y and all the staff members.

Department of Farmacy:

López E, Tuset M and all the staff members.

Department of Autoimmune diseases:

Espigol G, Espinona G and all the staff members.

## Funding

There are none funding or conflicts of interest to declare.

## Abbreviations

ALS: Advanced Life Support
ARDS: Acute Respiratory Distress Syndrome
CRP: C-Reactive Protein
ICU: Intensive Care Unit
MV: mechanical ventilation
PCR: Polymerase Chain Reaction
PS: Propensity Score
SD: Standard Deviation

## Notes

### Competing Interest Statement

The authors have declared no competing interest.

### Funding Statement

No funding to declare

### Author Declarations

The Institutional Ethics Committee of the Hospital Clinic of Barcelona approved the study and due to the nature of retrospective chart review, waived the need for inform consent from individual patients (Comite Etic Investigacio Clinica; HCB/2020/0273).

